# Non-COVID-19 deaths in the United States during the imposition of sheltering-in-place

**DOI:** 10.1101/2020.07.26.20162396

**Authors:** Ralph Catalano, Joan A. Casey, Alison Gemmill, Tim A. Bruckner

## Abstract

Public debate over imposed sheltering-in-place in the United States (US) includes the claim that non-COVID-19 deaths increased above those expected from history and from such deaths in Sweden. We test this claim by applying Box-Jenkins transfer function modeling to weekly deaths between December 29, 2013 and May 16, 2020 in the US and Sweden. In the eight weeks of imposed sheltering-in-place in the US – assumed begun in the week of March 22 – an average of 715 fewer non-COVID-19 deaths (95% CI: −1428, −2) occurred per week implying a total of 5720 less than expected from those in Sweden and from history. Results changed little (i.e., 5472 fewer deaths than expected) when we excluded deaths in New York City. Contrary to speculation, non-COVID-19 mortality appeared to decline in the US after shelter-in-place orders.

## Introduction

Popular^1^ and more scholarly^2^ media report that deaths of all kinds increased above expected levels in the United States (US) after the imposition of sheltering-in-place in March, 2020. This increase presumably included deaths due not only to Coronavirus Disease 2019 (COVID-19), but also to overwhelmed health care providers unable to treat persons emergently ill from other causes or reluctant to seek timely care. Citing these reports, commentators, including the President of the United States, have argued that stay-at-home orders and their sequelae produced the same or more deaths than would have occurred had states pursued the policy of voluntary distancing adopted by, for example, Sweden.^3-5^ Assessing the accuracy of such assertions would seem important because they influence individual and institutional responses to the pandemic. The scholarly literature, however, includes no attempts to determine whether the observed incidence of non-COVID-19 deaths in the US after the shelter-in-place orders of mid-March, and before their loosening in early to mid-May, rose above that expected from earlier trends and from the incidence of non-COVID-19 deaths in places, such as Sweden, that did not impose stay-at-home policies. We use rigorous time-series methods and the most current data to test the hypothesis that non-COVID-19 deaths in the US rose above levels expected from trends and from those in Sweden during the imposition of stay-at-home orders.

## Methods

We extracted, from the Human Mortality Database’s Short-term Mortality Fluctuations data series,^6^ weekly counts of deaths for the 332 weeks beginning December 29, 2013 and ending May 16, 2020. We ended our test period in mid-May for two reasons. First, many states began loosening restrictions by that time^7^ thereby ending the effects of imposed isolation. Second, although the most recent data at the time of our test (i.e., July 21, 2020) ended June 13, 2020, curators of the data characterize the most recent four weeks as preliminary.

We also obtained daily counts of COVID-19 deaths from the Your World in Data Website and aggregated them to weeks.^8^ We created our test variables by subtracting COVID-19 weekly deaths from all weekly deaths for both Sweden and the United States as well as, for reasons noted below, New York City.

Our tests used Box-Jenkins methods^9^ to implement the causal reasoning of Granger.^10-12^ These methods remove from weekly US deaths the variation predictable not only from deaths in Sweden, but also from autocorrelation (i.e., trends, seasonality, or the tendency to remain elevated or depressed after high or low values) observed in US deaths before social distancing. This approach controls “third variables” affecting both the US and Sweden as well as those unique to the US that exhibit autocorrelation. The approach also assures a statistically efficient estimation of coefficients because the adjusted series will meet the assumption of constant mean and serial independence.

We set the date of imposed isolation in the United States at the week starting March 22 when California, New York, and other large states issued stay-at-home orders and when cell phone data indicated most Americans started sheltering in place.^13^

Our test proceeded through the following steps.

1. We regressed the weekly number of non-COVID-19 deaths in the US on those in Sweden for the 324 weeks beginning December 29, 2013 and ending March 21, 2020. We specified Swedish deaths in the same week as, and one week earlier than, US deaths to ensure that Monday versus Sunday week starts did not affect results.
2. We used Box and Jenkins methods to detect and model autocorrelation including trends, cycles (e.g. seasonality), and/or the tendency to remain temporarily elevated or depressed after high or low values in the residuals of the regression estimated in step one.
3. We created a binary imposed-isolation variable scored one for the eight weeks starting March 22, 2020 and zero otherwise.
4. We estimated a Box-Jenkins transfer function that estimated non-COVID-19 deaths in the US from those in Sweden, the imposed-isolation variable described in step three, and parameters specifying autocorrelation detected in step two. We estimated this transfer function for the 332 weeks beginning December 29, 2013 and ending May 16, 2020. We inferred support for the hypothesized increase in non-COVID-19 deaths if the coefficient for the social distancing variable were positively signed and at least twice its standard error.

## Results

Non-COVID-19 deaths in the US over our 332 test weeks ranged from 46 388 to 67 491 with a mean of 53 229. Figure 1 shows these counts plotted over the test weeks. Weekly non-Covid deaths in Sweden averaged 1709 and ranged from 1441 to 2204.

**Figure 1.**
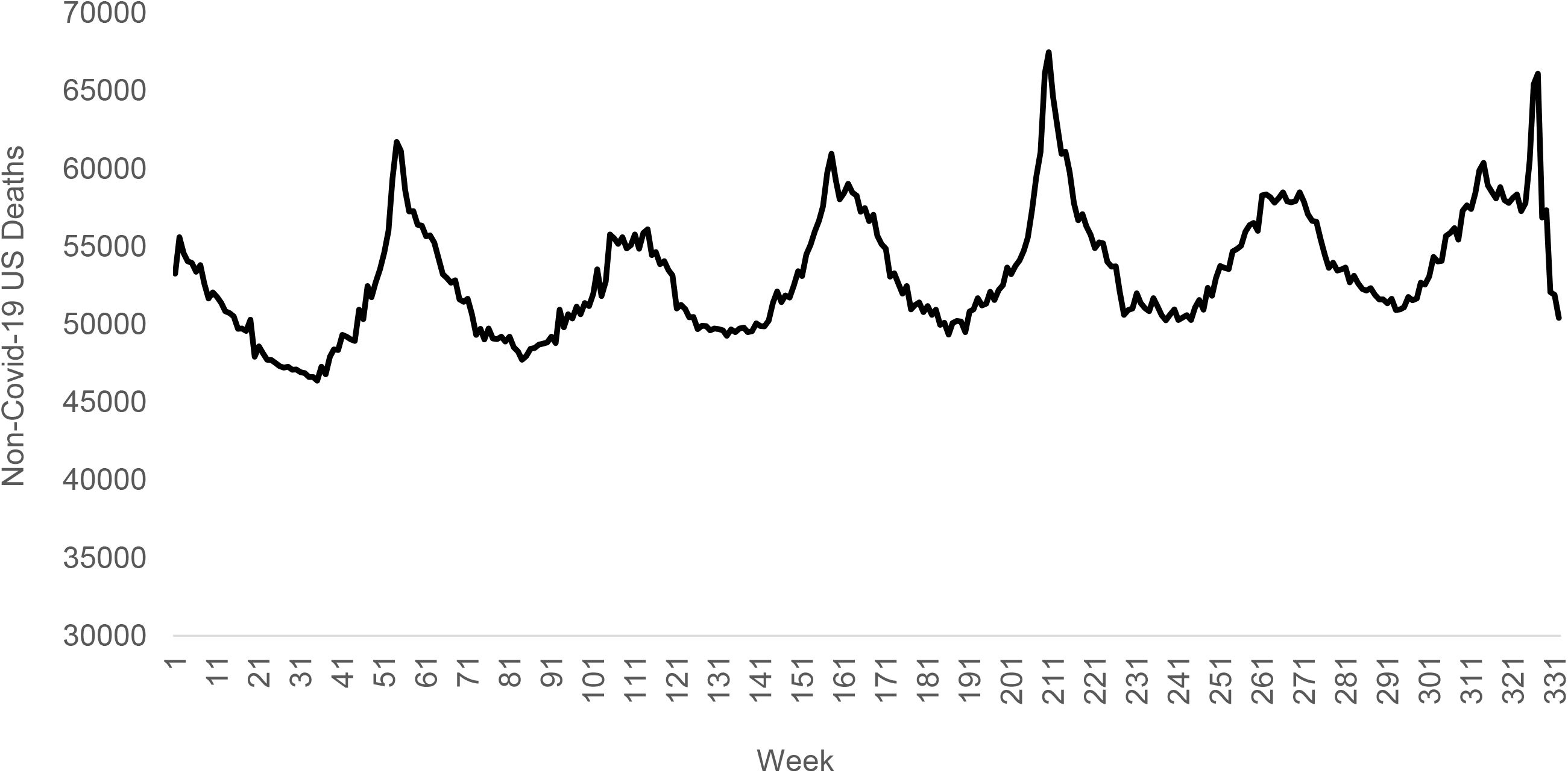
Counts of non-Covid-19 deaths in the US for the 332 weeks beginning December 29, 2013 and ending May 16,

The following equation shows the results, estimated in Step four, of our test.

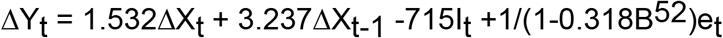

In which Y_t_ is the number of non-COVID-19 deaths in the US in week t. X_t_ and X_t-1_ are non-COVID-19 deaths in Sweden for weeks t and t-1. I_t_ is the imposed-isolation variable scored one for the eight weeks starting March 22, 2020 and zero otherwise. e_t_ is the error term for week t. Δ is the difference operator indicating that both X and Y had been differenced (i.e., value at t-1 subtracted from that at time t). B52 is the backshift operator or value of e at t-52 weeks. The coefficient estimating post March 22 change in non-COVID-19 deaths (i.e., −715) had a standard error of 363. The standard errors for the parameters expressing association with deaths in Sweden (i.e., 1.758 and 3.845) were 0.747 and 0.738 respectively. The autoregressive parameter at t-52 (i.e, 0.368) gauges seasonality and had a standard error of 0.066.

Contrary the hypothesized increase in non-COVID-19 deaths, the negatively signed coefficient for the imposed-isolation variable (i.e., −715, 95% CI: −1428, −2), implies a net *decrease* over the eight weeks following sheltering in place. Applied across the eight weeks, the coefficient suggests about 5720 fewer than expected deaths.

Figure 2 shows our results more graphically by plotting the differences between observed non-COVID-19 deaths in the US for the last 52 weeks of the test period and their corresponding counterfactuals (i.e., values expected from autocorrelation and from non-COVID-19 deaths in Sweden). Plotting only the last year of weekly differences allows closer inspection of the critical data than would plotting all 332 weeks. The dashed lines show the 95% confidence interval and the points show the differences. Consistent with reports noted at the outset of this article, non-COVID-19 deaths rose above expected in the two weeks after imposed isolation. Such deaths, however, then fell below expected.

**Figure 2.**
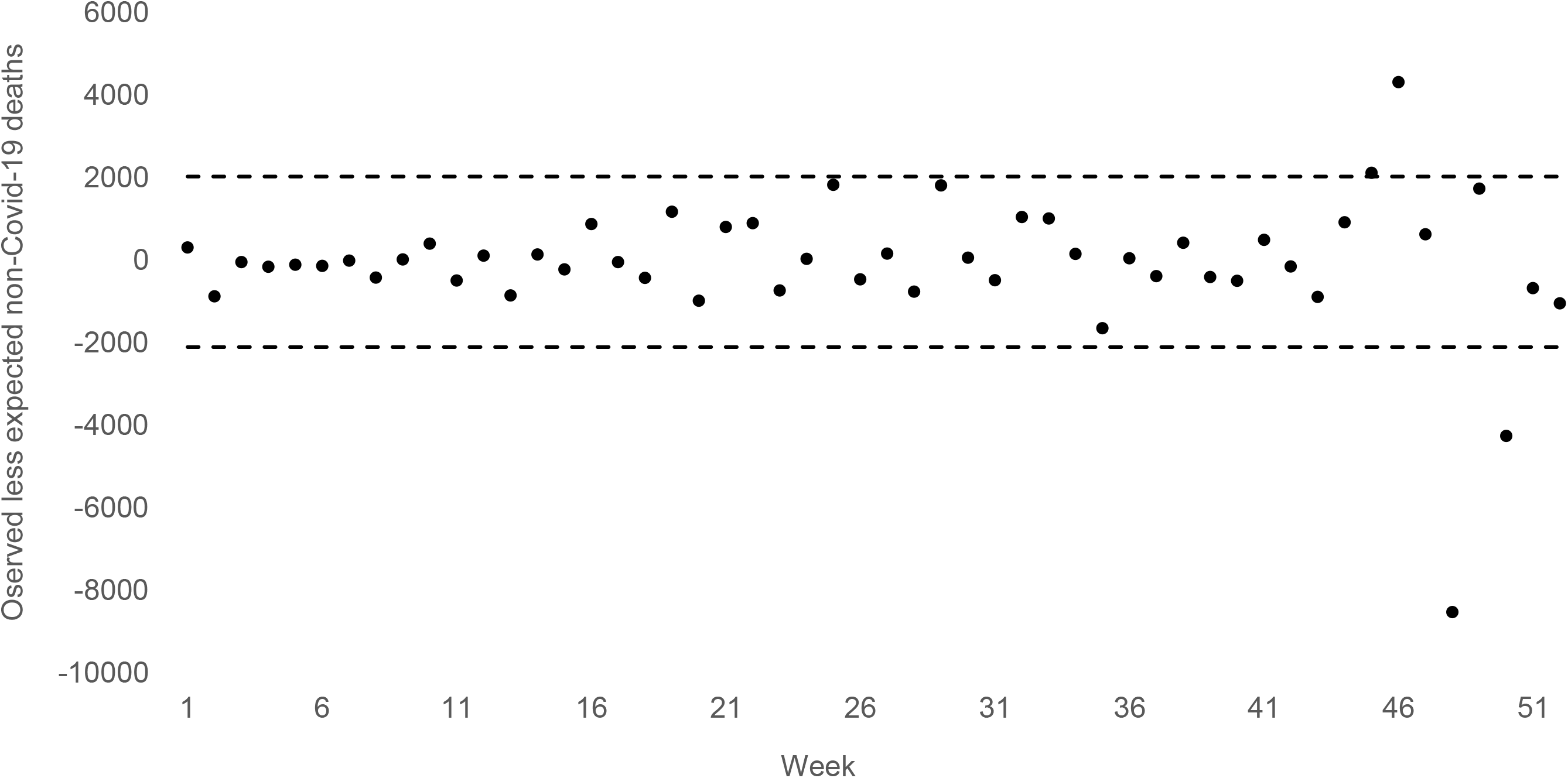
Observed less expected non-COVID-19 deaths in the United States and their 95% confidence interval for the 52 weeks starting 5/20/2019 and ending 5/17/2020. Expected values derived from non-COVID-19 deaths in Sweden and from autocorrelation for 324 weeks before the imposition of social distancing.

Anticipating curiosity over how the overwhelming of health care facilities in New York City influenced our findings, we repeated our test without non-COVID-19 deaths reported for the city. Results were essentially the same in that we found a negatively signed coefficient for imposed isolation (i.e., −684; SE = 346) that implied about 5472 fewer than expected deaths in the eight weeks.

We also anticipate the question of how the Swedish death covariate affected our results. We repeated our test but estimated counterfactuals from only autocorrelation in US non-COVID-19 deaths before March 2020. Results did not support the hypothesized increase in these deaths during imposed isolation. As with the main test, we observed fewer (i.e., −5840) than expected non-Covid-19 deaths in eight weeks of imposed isolation.

## Discussion

Our results do not support the argument that imposed isolation in the US produced the same or more deaths than would have occurred had states pursued less coercive policies like those adopted by Sweden. We found approximately 5720 fewer non-COVID-19 deaths than expected from history and from similar deaths in Sweden during the two months following imposition of isolation.

Strengths of our test include that no approach better implements Granger Causation than does the Box-Jenkins method we employed, and that we used the most reliable data available at the time of our test. Weaknesses include that the data do not provide enough detail to identify the causes of death that fell below expected levels. We further acknowledge that our results do not describe the sequelae of the “loosening” of shelter-in-place restrictions after our test period.

## Data Availability

All data available at Human Mortality Database and Our World in Data

https://www.mortality.org/

https://ourworldindata.org/

